# Chronic Kidney Disease of Unknown Etiology (CKDu) as an Underappreciated Cause of Emergent Hemodialysis Utilization in the United States

**DOI:** 10.1101/2025.02.28.25323090

**Authors:** Anna Strasma, Matthew R. Sinclair, Lawrence P. Park, Hal H. Zhang, Sreedhar A. Mandayam, Maulin Kirit Shah, Christina M. Wyatt, Rebecca S.B. Fischer

## Abstract

**Introduction:** End stage kidney disease (ESKD) affects an estimated 5500 persons living in the United States without legal residency documentation. One likely, but underappreciated cause of ESKD in the Hispanic migrant population, is chronic kidney disease of unknown etiology (CKDu). CKDu is an interstitial nephritis that disproportionately affects young adult agricultural workers in Central America who lack traditional risk factors for kidney disease. In underserved populations, such as those at risk for CKDu, substantial barriers to optimal kidney care translate to poorer health outcomes and widening health disparities. Without funding for non-emergent healthcare, this underserved population, often only have access to hemodialysis (HD) once a life-threatening condition occurs. Despite the presence of a migrant population from CKDu endemic countries and anecdotes of its presence, CKDu has very rarely been directly investigated or documented in the US. We undertook this study to establish the existence of CKDu in the United States and to characterize CKDu as a cause of ESKD in patients accessing emergent HD.

**Methods:** In a retrospective cross-sectional study among patients receiving emergent HD in Texas, we analyzed medical record data from a large, county hospital. We ascertained cause of ESKD and underlying hypertension and diabetes and compared these proportions to data on patients on maintenance HD from the US Renal Data System (USRDS). Undocumented immigrants are largely excluded from the USRDS, as with many health statistics databases in the US. We identified patients whose clinicians had indicated CKDu as a diagnosis and classified others as having suspected CKDu or possible CKDu based on clinically informed criteria.

**Results:** We identified 346 patients with ESKD requiring emergent HD (2012-2015), who were younger than patients in the USRDS (median age 52 yrs vs. 61 yrs, p <0.001), had more comorbid diabetes (60% vs. 47%, p <0.001), and more often had an unknown cause of ESKD (16% vs. 4%, p<0.001). Patients requiring emergent HD also had less frequent arteriovenous access (12% vs. 82%, p<0.001). ESKD attributed to diabetes and/or hypertension accounted for only 67% of emergent HD patients, compared to 81% of USRDS patients (p<0.001). 14% of the patients on emergent HD died during the study period. Four patients had been clinically diagnosed with CKDu, while we classified 14 with suspected CKDu and 40 with possible CKDu, for a total of 58 patients (17%) with potentially CKDu-related ESKD.

**Conclusion:** Our analysis suggests that up to 17% of patients in this population utilizing emergent HD had CKDu-related ESKD, suggesting that CKDu is likely underdiagnosed in the US. Further, patients receiving emergent HD were younger but were at higher risk of infection or complication than patients receiving scheduled, maintenance HD. Understanding CKDu and improving access to scheduled dialysis for migrants without legal residency documentation should be prioritized to reduce stress on the healthcare system and improve health among vulnerable populations in the US.

## INTRODUCTION

It is well established that health and healthcare inequities plague the estimated 11 million individuals who lack legal residency documentation in the United States, most of whom are from Mexico and Central America.^1^ Migrant populations often have complex health determinants, such as instability in their countries of origin, disruption of social networks, discrimination, limited healthcare access, and language barriers.^2^ For the undocumented immigrant who has or who develops a chronic health condition, such as kidney disease, barriers to accessing healthcare serve to further drive health disadvantages. These barriers include, among other things, distrust in the healthcare system, difficulty accessing and navigating healthcare due to cultural differences and language barriers, and limited resources to obtain essential care. In particular, many go without healthcare because of economic considerations, such as public health insurance eligibility restrictions, lack of employer-provided healthcare, and inability to afford private insurance.^2^

End stage kidney disease (ESKD) is a chronic condition wherein the kidneys have such poor function that dialysis or a kidney transplant is needed for mere survival. It requires substantial medical management and has various systemic health effects. An estimated 5,500 – 9,000 US immigrants without legal residency documentation have ESKD.^3^ In the US, Medicare provides most citizens or permanent residents with several ESKD therapeutic options, including thrice weekly scheduled outpatient hemodialysis (HD) for health maintenance for the duration of life.^4^ However, most immigrants without documentation lack access to this benefit.^5^ In such cases, patients may have access to HD only once a life-threating condition occurs, also called “emergent HD”.^3,6^ Emergent HD is possible once patients develop serious symptoms, such as shortness of breath or high serum potassium levels, when emergency departments are obligated to provide stabilizing treatment under the Emergency Medical Treatment and Labor Act.^3^ However, because patients with ESKD have a chronic, life-threatening disease that requires routine management, they will invariably require subsequent HD, often within several days. Because emergent HD is only available after a threshold of disease severity is crossed, these patients tend to be much sicker. They have higher mortality rates and higher per-patient healthcare costs than patients receiving scheduled maintenance HD.^7,8^

An underlying feature to patients requiring emergent HD may be chronic kidney disease (CKD) of unknown etiology (CKDu), a kidney disease endemic to South Asia and Central America and occasionally reported elsewhere.^9^ CKDu occurs in the absence of traditional kidney disease antecedents, such as diabetes or hypertension, with a younger onset than is observed in other CKD entities. Patients are largely thought to be asymptomatic in the early stages, during which time their kidney disease goes undetected, and they remain undiagnosed until they develop advanced disease. Kidney biopsies, though rarely performed for CKDu, demonstrate interstitial nephritis.^9–11^ While the cause is unknown, it is associated with heat exposure, agricultural labor, and exposure to environmental toxicants.^5^ Mostly characterized in young, male agricultural workers in Central America, CKDu is remarkably common in certain regions.^5^ In Tierra Blanca, Mexico, CKD prevalence is estimated at 25%, with over half of cases occurring due to unknown etiology.^12^ In El Salvador, CKD is the second leading cause of death, and CKDu prevalence is 6% among males in that country.^13^ There are limited primary prevention strategies or treatment for CKDu, and specialized kidney care and interventions such as HD or transplant are often unavailable or difficult to access in areas heavily burdened with CKDu.^14^ Hence, progression to ESKD and kidney-related death is common for many individuals living with CKDu outside of the US.

Within the US, CKDu has scarcely been considered and has only been indirectly studied.^15,16^ Most knowledge about the disease and its origins stems from research conducted in established communities in Central America where CKDu has emerged and become endemic.^15^ From anecdotes and ecological evidence, CKDu is part of the kidney disease landscape in the US, the extent of which remains unknown. Some US agriculture workers, including migrants from Mexico and Central America, have biomarkers consistent with suspected CKDu etiologies. ^17–21^ The United States Renal Data System (USRDS), which collates information about ESKD in the US, reflects that Hispanic patients initiate dialysis at younger ages than their non-Hispanic white counterparts and have a higher proportion of unexplained kidney disease, although CKDu is not specifically parsed out in that data.^15^ Undocumented individuals are rarely represented in the USRDS, as in other sources of national health statistics, which means the true burden of unexplained kidney disease is likely much higher than current estimates, especially in that special population.^22,23^ Despite the systematic underrepresentation of undocumented immigrants in the USRDS, two ESKD hotspots emerged from that data – the agricultural regions of the San Joaquin Valley in California and the Rio Grande Valley in Texas – where the population is majority Hispanic.^16^ It is yet unknown whether CKDu occurs de novo in the US, even among populations that could be considered at high risk. Although CKDu has not yet been directly documented in the US, existing data support the suspicions that it is present at some level.^15,16,24^

In a setting where a majority of undocumented individuals in the US are from CKDu-endemic areas in Latin America, we hypothesize that a notable proportion of ESKD in the US is attributable to CKDu, resulting in a disproportionate burden of CKDu among patients requiring emergent HD. By measuring the prevalence of patients with CKDu-related ESKD in this patient population, we can gain insight into the emergence of CKDu in the US, with the goal of improving early disease detection and raising awareness among primary care clinicians and nephrologists alike. This could help develop strategies to better recognize and characterize individuals with CKDu, while also recognizing risk factors for its development to prevent disease progression. Furthermore, by better understanding the causes of CKDu and progression, this valuable information could be shared with physicians caring for these patients in endemic areas.

In this retrospective, cross-sectional analysis of patients accessing emergent HD at a large public hospital in Harris County, Texas, we sought to demonstrate that CKDu is a disease entity presenting at the clinical interface in the US and to provide insight into CKDu as a potential contributor to emergent HD utilization in Texas.

## METHODS

### Study setting

Harris County, Texas cares for its uninsured and indigent population through a large, public healthcare system in which 25% of the patient population is comprised of undocumented immigrants.^25^ The majority (85%) of immigrants in the county originate from Mexico and Central America.^26^ Owed to existing healthcare infrastructure for uninsured individuals in Harris County, outpatient maintenance HD during the study period was available to some, but not all, undocumented immigrants, leaving an estimated 32% with emergent HD as their only option.^27,28^

### Data collection

We conducted a cross-sectional secondary data analysis using retrospective medical record data gathered on patients who received emergent HD during 2012-2015, primarily through the emergency department.^29^ Patients were not included in the analysis if they were simultaneously receiving maintenance HD at another site or if they had received HD for fewer than 30 days in the healthcare system or fewer than 5 times during the study period. The parent study was reviewed and approved by the Institutional Review Board (IRB) of Baylor College of Medicine, and this analysis was approved by Duke University Health System IRB.^29^

Data on demographics, comorbidities, HD access, cause of ESKD, history of kidney biopsy, duration and frequency of emergency-only HD, catheter-related blood stream infections, and mortality were collected through medical record review and available for analysis. Dialysis during hospital admissions and time without HD for >30 days were not included since the patient may have received HD in another system.

### Data analysis

We compared the Harris County emergent HD study population to the United States Renal Data System (USRDS), which collects, analyzes, and distributes information about ESKD in the US.^23^ We adopted the USRD classification system on the cause of ESKD and applied it to our study population. In our study population, we classified participants who lacked a specified ESKD etiology according to clinically informed CKDu case definitions. Patients were categorized as having *clinically diagnosed CKDu* if recorded as such in their clinical record.

Based on commonly described features of CKDu patients, we defined *suspected CKDu* as young (18-44 years old), male, and non-diabetic with an unknown cause of ESKD.^30–32^ Since CKDu is not restricted by sex, age, and occurs in other patient groups, including patients with interstitial nephritis (the nonspecific pathologic feature of CKDu) or hypertension (as it is uncommon for hypertension alone to cause ESKD at young ages^33,34^), we broadly classified patients with unknown ESKD cause from these other groups as *possible CKDu*. ^9,10,32,35^ Individuals with diabetes were excluded from all CKDu groups. All participants in the *clinically diagnosed, suspected,* and *possible* CKDu will collectively be referred to as have potential CKDu-related ESKD.

Patients receiving emergent HD for ESKD due to diabetic nephropathy were compared to those with CKDu. Both groups required emergent HD, which is associated with poor outcomes. The diabetic nephropathy group was selected as a comparison because it represents a traditional risk factor absent from CKDu. There was no overlap between the groups, as CKDu classification requires the absence of diabetes. Due to the small number of *clinically diagnosed* CKDu cases, these were combined with *suspected* CKDu cases for analysis. *Possible* CKDu cases were not included to limit misclassification. Continuous variables are described with medians and quartiles (median [IQR]). Categorical variables are described in frequencies and proportions (n [%]). We compared characteristics of our emergent HD study population and the USRDS data using non-parametric tests (Kruskal Wallis test and Fisher Exact Test), considering differences statistically significant at p<0.05. Analyses were done in R 4.2.1 (R Foundation for Statistical Computing, Vienna, Austria).^36^

## RESULTS

### Characterizing patients receiving emergent HD

We compared the characteristics of 346 subjects receiving emergent HD to the USRDS reference population (n=42,243) of patients receiving scheduled maintenance HD in Texas during the same time period **(Table 1).**^37^ Patients receiving emergent HD were younger (median age 51.5 yrs vs 61.0 yrs; p<0.001) and more often had diabetes (60.1% vs 47%; p<0.001) than the reference group. They less frequently had arteriovenous (AV) access, such as an AV fistula or graft (11.6% vs 82% in the reference group; p< 0.001) and more often had central venous catheters (CVC) for HD (93.3% vs 18%, respectively; p< 0.001). In the study population, emergent HD was utilized 5 times monthly (median; [IQR 4-7]). During the 3-year period we reviewed, 49 individuals (14.2%) died or transitioned to palliative care. Patients who died were significantly older than those who survived (median 60.0 yrs [IQR 52-70] vs 49.5 yrs [IQR 40-59]; p<0.001).

**Table 1:**
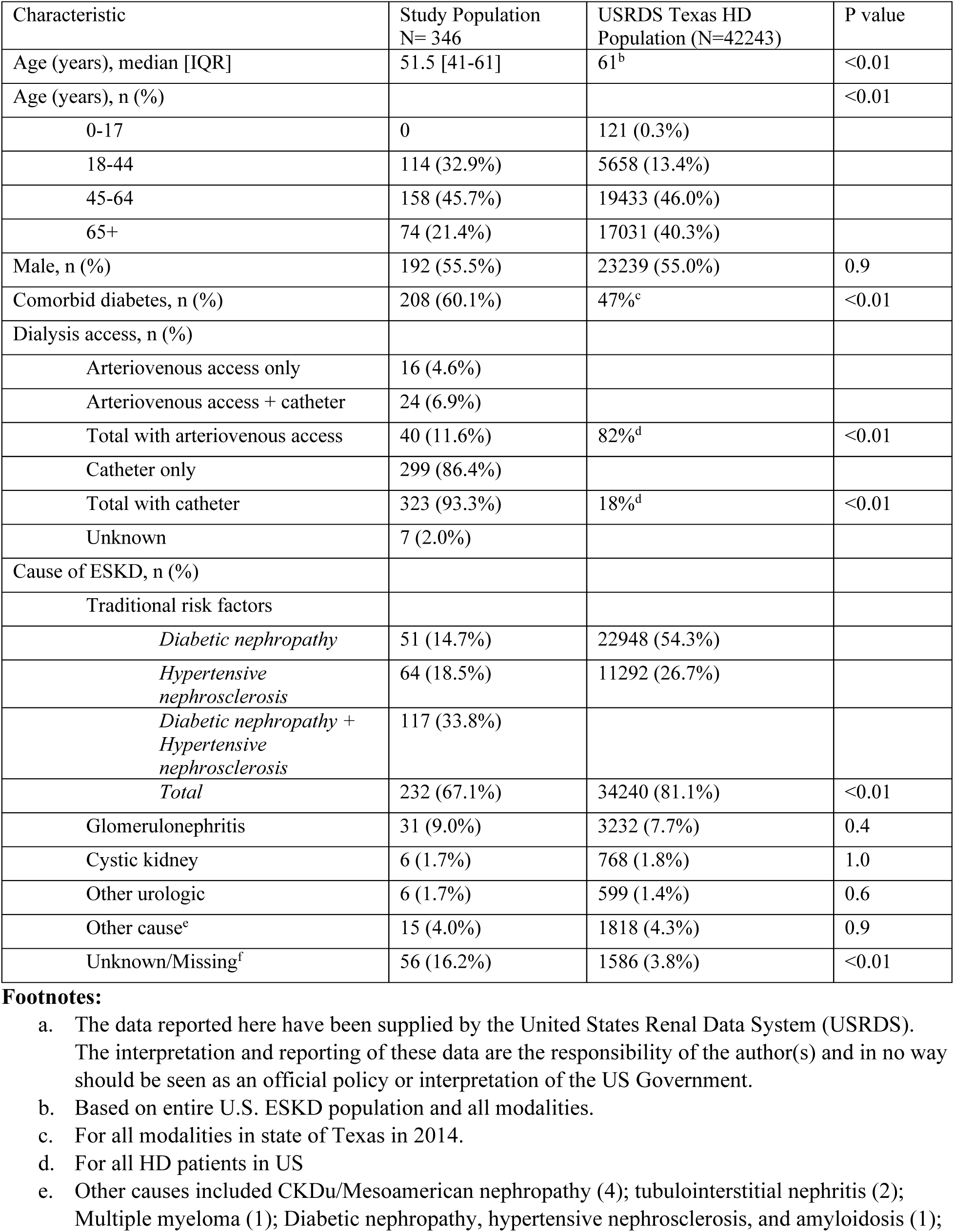
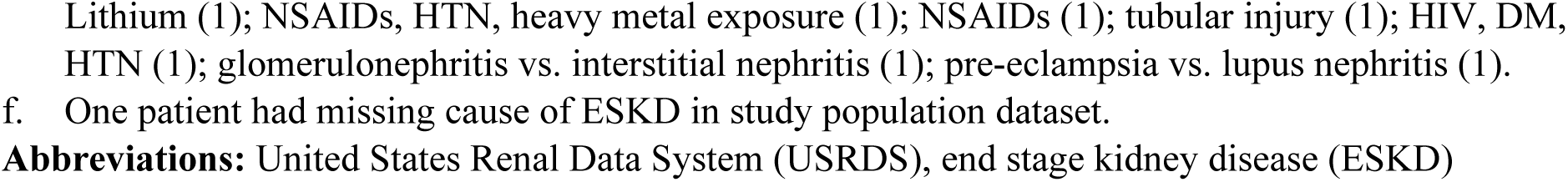
Characteristics of the emergent HD study population and the HD population in the USRDS for Texas (2014)^a^.

| Characteristic | Study Population<br>N= 346 | USRDS Texas HD<br>Population (N=42243) | P value |
| --- | --- | --- | --- |
| Age (years), median [IQR] | 51.5 [41-61] | 61<sup>b</sup> | <0.01 |
| Age (years), n (%) |  |  | <0.01 |
| 0-17 | 0 | 121 (0.3%) |  |
| 18-44 | 114 (32.9%) | 5658 (13.4%) |  |
| 45-64 | 158 (45.7%) | 19433 (46.0%) |  |
| 65+ | 74 (21.4%) | 17031 (40.3%) |  |
| Male, n (%) | 192 (55.5%) | 23239 (55.0%) | 0.9 |
| Comorbid diabetes, n (%) | 208 (60.1%) | 47%<sup>c</sup> | <0.01 |
| Dialysis access, n (%) |  |  |  |
| Arteriovenous access only | 16 (4.6%) |  |  |
| Arteriovenous access + catheter | 24 (6.9%) |  |  |
| Total with arteriovenous access | 40 (11.6%) | 82%<sup>d</sup> | <0.01 |
| Catheter only | 299 (86.4%) |  |  |
| Total with catheter | 323 (93.3%) | 18%<sup>d</sup> | <0.01 |
| Unknown | 7 (2.0%) |  |  |
| Cause of ESKD, n (%) |  |  |  |
| Traditional risk factors |  |  |  |
| *Diabetic nephropathy* | 51 (14.7%) | 22948 (54.3%) |  |
| *Hypertensive nephrosclerosis* | 64 (18.5%) | 11292 (26.7%) |  |
| *Diabetic nephropathy + Hypertensive nephrosclerosis* | 117 (33.8%) |  |  |
| *Total* | 232 (67.1%) | 34240 (81.1%) | <0.01 |
| Glomerulonephritis | 31 (9.0%) | 3232 (7.7%) | 0.4 |
| Cystic kidney | 6 (1.7%) | 768 (1.8%) | 1.0 |
| Other urologic | 6 (1.7%) | 599 (1.4%) | 0.6 |
| Other cause<sup>e</sup> | 15 (4.0%) | 1818 (4.3%) | 0.9 |
| Unknown/Missing<sup>f</sup> | 56 (16.2%) | 1586 (3.8%) | <0.01 |

- a. The data reported here have been supplied by the United States Renal Data System (USRDS). The interpretation and reporting of these data are the responsibility of the author(s) and in no way should be seen as an official policy or interpretation of the US Government. - b. Based on entire U.S. ESKD population and all modalities. - c. For all modalities in state of Texas in 2014. - d. For all HD patients in US - e. Other causes included CKDu/Mesoamerican nephropathy (4); tubulointerstitial nephritis (2); Multiple myeloma (1); Diabetic nephropathy, hypertensive nephrosclerosis, and amyloidosis (1);
Lithium (1); NSAIDs, HTN, heavy metal exposure (1); NSAIDs (1); tubular injury (1); HIV, DM, HTN (1); glomerulonephritis vs. interstitial nephritis (1); pre-eclampsia vs. lupus nephritis (1). f. One patient had missing cause of ESKD in study population dataset.
**Abbreviations:** United States Renal Data System (USRDS), end stage kidney disease (ESKD)

The most common recorded causes of ESKD in the emergent HD group were diabetic nephropathy (14.7%), hypertensive nephrosclerosis (18.5%), or both (33.8%), together accounting for 67.1% of emergent HD patients, while these accounted for a total of 81.1% in the reference group (p< 0.001; **Table 1**). Notably, 16.2% of emergent HD patients lacked an attributable cause of ESKD, versus only 3.8% of maintenance HD patients (p<0.001). In the emergent group, 10.1% underwent kidney biopsy.

### Estimated prevalence of CKDu in emergent HD patients

Four participants (1.1%) in the emergent HD study population were *clinically diagnosed* as having ESKD from CKDu **(Table 2)**; none had undergone kidney biopsy. In addition to these 4 diagnosed CKDu cases, 14 individuals (4.0%) had *suspected* CKDu, and an additional 40 cases were classified as *possible* CKDu (11.6%). Thus, the prevalence of potential CKDu-related ESKD in the patients requiring emergent HD was 16.8%.

**Table 2:** Classification criteria for CKDu in the emergent HD study population (n=##)

| CKDu Classification | n (%) | Definition |
| --- | --- | --- |
| Clinically diagnosed | 4 (1.1%) | Patients determined by their clinicians to have CKDu as the most likely cause of ESKD |
| Suspected | 14 (4.0%) | Males from 18-44 years old with unknown cause of ESKD and without comorbid diabetes |
| Possible | 40 (11.6%) | Patients 18-44 years old without comorbid diabetes who are:<br>(a) Females with unknown cause of ESKD (n= 6) **or**<br>(b) Males or females with chronic interstitial nephritis (n= 2) **or**<br>(c) Males or females with hypertension as cause of ESKD (n= 32) |
| **Total** | **58 (16.8%)** |  |

**Abbreviations:** Chronic Kidney Disease of Unknown Etiology (CKDu), End stage kidney disease (ESKD)

### Characteristics of patients with CKDu

To further understand if CKDu patients differed from other ESKD patients utilizing emergent HD in our study population, we compared the subset of patients with *confirmed* and *suspected* CKDu (CKDu group; n=18) to the subset of patients with diabetic nephropathy (DN group, n=51) (**Table 3**). The CKDu group was younger (32.5 yrs [IQR 27-39]) than the DN group (54.0 yrs [IQR 47-64]; p=0.041) and was more likely to have AV access (33.3% vs 5.9%, p=0.008). In the DN group, 13 participants (25.5%) died or went to palliative care, compared to none in the CKDu group (p = 0.015).

**Table 3:** Characteristics of the CKDu group and the DN group in the emergent HD study population.

|  | Diabetic nephropathy (n = 51) | CKDu diagnosed or suspected (n=18) | P value |
| --- | --- | --- | --- |
| **Demographics** |  |  |  |
| Age, median (years) [quartiles] | 54 [47-64] | 32.5 [27-39] | 0.04 |
| Males, n (%) | 26 (51.0%) | 18 (100%)<sup>b</sup> | <0.01 |
| Dialysis access |  |  |  |
| *Total with arteriovenous access* | 3 (5.9%) | 6 (33.3%) | 0.01 |
| *Total with catheter* | 50 (98.0%) | 17 (94.4%) | 0.5 |
| Frequency of dialysis per month, median [quartiles] | 5 [4-6] | 6 [5-7] | 0.02 |
| **Outcomes** |  |  |  |
| Catheter related blood stream infections, n (%) | 15 (29.4%) | 4 (22.2%) | 0.8 |
| Drug resistant infections<sup>a</sup> | 2 (3.9%) | 0 |  |
| Death or palliative care, n (%) | 13 (25.5%) | 0 | 0.02 |

**Footnote:** a. All drug-resistant infections were methicillin-resistant *Staphylococcus aureus* infections. b. CKDu diagnosed or suspected classification was limited to males.
**Abbreviations:** Chronic Kidney Disease of Unknown Etiology (CKDu); diabetic nephropathy (DN)

## DISCUSSION

ESKD is a substantial and challenging chronic health condition for patients and their families, as well as a growing issue in migrant health. We highlight CKDu as an underrecognized cause of ESKD and emergent HD in the US, based on evidence from a public healthcare system in a major medical center in Texas. Although we found that CKDu was an infrequently recorded clinical diagnosis, the proportion of ESKD patients undergoing emergent HD who met a CKDu clinico-epidemiologic profile was not inconsequential, as high as an estimated 17%. As the epidemics of CKDu surge around the world, an opportunity to raise clinical and public health awareness in the US provides hope for early disease detection and timely intervention. With improved awareness, counseling, and management of risk factors, CKD progression to ESKD may be mitigated both in the US and abroad.

Patients undergoing emergent HD in this system, which cares for many undocumented immigrants from CKDu endemic countries in Latin America, have important differences from patients utilizing scheduled maintenance HD. In our study, they were younger, and, despite diabetes being a common comorbidity, diabetic nephropathy was infrequently implicated as their cause of ESKD; rather, the high proportion of diabetic patients in our study likely reflects the high burden of diabetes among the US Hispanic population.^38,39^ Further, patients characterized as having CKDu – or who lack a clinical diagnosis but fit the clinical and epidemiologic profile of disease – were younger still than other emergent HD patients, receiving HD in their second and third decades of life, remarkably early even for this disease that notoriously appears in young adulthood.^40,41^ This special population of young patients from CKDu high-risk settings may benefit substantially from targeted approaches to early kidney screening and improved care to avoid an otherwise disproportionate number of future disability-adjusted life years and the premature mortality associated with CKDu.

Migrants undergo barriers to routine, quality medical care. Emergent HD patients were less likely to have undergone kidney biopsy, perhaps one reason they more often lacked a definitive ESKD etiology. Emergent HD care is not provided at the standard offered by outpatient maintenance HD. In our study population, HD was only performed a median of 5 times monthly compared to the standard 3 times weekly (13 times monthly). This can be attributed to requiring laboratory abnormalities or symptoms to be an emergency before dialysis is performed. They also have higher use of CVCs and less AV access, which may be due to elective AV access placement requiring additional outpatient care and multiple visits to both a nephrologist and a surgeon, compared to tunneled dialysis catheter placement, which often occurs in the hospital. CVCs have been shown to increase infection risk and mortality compared to AV access. However, most emergent HD patients with AV access also had a CVC, so there was similar prevalence of CVCs between the two groups, as was the incidence of catheter related blood stream infections. Unfortunately, mortality was high in our study group, with 14% experiencing either death or transition to palliative care within the three-year study period. Mortality is likely underestimated due to loss of follow-up and because palliative care was not specifically queried for each participant.

### Limitations

The nature of this secondary analysis of data retrieved through medical record abstraction limits our ability to address important questions that cannot be queried from the original data (e.g., birth country) and introduces the possibility of misclassification, a common problem with retrospective medical record data. However, based on our team’s own experience at this clinical site, we feel the data fairly represents our own observations. Similarly, mortality and etiology may be incompletely captured in the records, particularly if patients sometimes seek care elsewhere. The public healthcare system in Harris County serves a large population of migrants, most from Latin America, and at the time of this study provided maintenance HD to approximately one third of undocumented ESKD patients. Therefore, patients in our analysis may not represent all emergent HD patients in this setting or others. Replicating our findings in other settings, including through active surveillance, will be important. Race-ethnicity was not a variable available to us in this analysis; however, authors previously identified this patient population as 90% Hispanic, and emergent HD patients consist primarily of undocumented immigrants.^29^

## CONCLUSION

We highlight CKDu as an underrecognized cause of ESKD and emergent HD utilization in the US. Endemic to Mexico and Central America, CKDu is the leading cause of death in some communities, making it likely that it would be observed in patients moving to or visiting the US.^42,43^ Any of these patients who lack legal residence documentation are subject to substandard dialysis care and outcomes, as emergent HD may be the only option for life-preserving therapy. In order to prevent this, clinical awareness in the primary care setting is needed, including where vulnerable undocumented patients normally seek care. Early screening, detection, referral to specialty care, and expanded access to scheduled outpatient dialysis and kidney transplantation would improve health outcomes for these members of our US population. Understanding more about CKDu in the US stands to advance and support scientific knowledge about the peculiar disease entity and its underlying cause(s). Recognizing and addressing CKDu in the US is crucial for promoting equitable healthcare access, improving patient outcomes, and advancing understanding of this global public health challenge.

## Data Availability

All data produced in the present study are available upon reasonable request to the authors.

## REFERENCES

1. Capps R, Gelatt J, Van Hook J, Fix M. Commentary on “The number of undocumented immigrants in the United States: Estimates based on demographic modeling with data from 1990-2016.” PLoS ONE. 2018;13(9):e0204199. doi:10.1371/journal.pone.0204199

2. Ornelas IJ, Yamanis TJ, Ruiz RA. The Health of Undocumented Latinx Immigrants: What We Know and Future Directions. Annu Rev Public Health. 2020;41:289–308. doi:10.1146/annurev-publhealth-040119-094211

3. Rodriguez R, Cervantes L, Raghavan R. Estimating the prevalence of undocumented immigrants with end-stage renal disease in the United States. Clin Nephrol. 2020;93(1):108–112. doi:10.5414/CNP92S119

4. LCD – Frequency of Hemodialysis (L34575). Accessed August 22, 2024. https://www.cms.gov/medicare-coverage-database/view/lcd.aspx?lcdId=34575&ver=43

5. Jayasumana C, Orantes C, Herrera R, et al. Chronic interstitial nephritis in agricultural communities: A worldwide epidemic with social, occupational and environmental determinants. Nephrol Dial Transplant. 2017;32(2):234–241. doi:10.1093/ndt/gfw346

6. Kuruvilla R, Raghavan R. Health care for undocumented immigrants in Texas: past, present, and future. Tex Med. 2014;110(7):e1.

7. Sheikh-Hamad D, Paiuk E, Wright AJ, Kleinmann C, Khosla U, Shandera WX. Care for immigrants with end-stage renal disease in Houston: a comparison of two practices. Tex Med. 2007;103(4):54–58, 53.

8. Cervantes L, Tuot D, Raghavan R, et al. Association of Emergency-Only vs Standard Hemodialysis With Mortality and Health Care Use Among Undocumented Immigrants With End-stage Renal Disease. JAMA Intern Med. 2018;178(2):188–195. doi:10.1001/jamainternmed.2017.7039

9. Wijkström J, Leiva R, Elinder CG, et al. Clinical and Pathological Characterization of Mesoamerican Nephropathy: A New Kidney Disease in Central America. Am J Kidney Dis. 2013;62(5):908–918. doi:10.1053/j.ajkd.2013.05.019

10. Fischer RSB, Vangala C, Truong L, et al. Early detection of acute tubulointerstitial nephritis in the genesis of Mesoamerican nephropathy. Kidney Int. 2018;93(3):681–690. doi:10.1016/j.kint.2017.09.012

11. Correa-Rotter R, García-Trabanino R. Mesoamerican Nephropathy. Semin Nephrol. 2019;39(3):263–271. doi:10.1016/j.semnephrol.2019.02.004

12. Aguilar DJ, Madero M. Other Potential CKD Hotspots in the World: The Cases of Mexico and the United States. Semin Nephrol. 2019;39(3):300–307. doi:10.1016/j.semnephrol.2019.02.008

13. Orantes-Navarro CM, Almaguer-López MM, Alonso-Galbán P, et al. The Chronic Kidney Disease Epidemic in El Salvador: A Cross-Sectional Study. MEDICC Rev. 2019;21:29–37.

14. Sanchez Polo V, Garcia-Trabanino R, Rodriguez G, Madero M. Mesoamerican Nephropathy (MeN): What We Know so Far. Int J Nephrol Renov Dis. 2020;13:261–272. doi:10.2147/IJNRD.S270709

15. Claudel SE, Chan M, Scammell MK, Waikar SS. Challenges and Opportunities: Studying CKDu in the United States. Kidney360. 2024;5(4):607. doi:10.34067/KID.0000000000000408

16. Bragg-Gresham J, Morgenstern H, Shahinian V, Robinson B, Abbott K, Saran R. An analysis of hot spots of ESRD in the United States: Potential presence of CKD of unknown origin in the USA? Clin Nephrol. 2020;93(1):113–119. doi:10.5414/CNP92S120

17. Mizelle E, Larson KL, Bolin LP, Kearney GD. Fluid Intake and Hydration Status Among North Carolina Farmworkers: A Mixed Methods Study. Workplace Health Saf. 2022;70(12):532–541. doi:10.1177/21650799221117273

18. Houser MC, Mac V, Smith DJ, et al. Inflammation-Related Factors Identified as Biomarkers of Dehydration and Subsequent Acute Kidney Injury in Agricultural Workers. Biol Res Nurs. 2021;23(4):676–688. doi:10.1177/10998004211016070

19. Chicas RC, Elon L, Xiuhtecutli N, et al. Longitudinal Renal Function Degradation Among Florida Agricultural Workers. J Occup Environ Med. 2024;66(9):694. doi:10.1097/JOM.0000000000003142

20. Mix J, Elon L, Vi Thien Mac V, et al. Hydration Status, Kidney Function, and Kidney Injury in Florida Agricultural Workers. J Occup Environ Med. 2018;60(5):e253. doi:10.1097/JOM.0000000000001261

21. Moyce S, Joseph J, Tancredi D, Mitchell D, Schenker M. Cumulative Incidence of Acute Kidney Injury in California’s Agricultural Workers. J Occup Environ Med. 2016;58(4):391–397.

22. Campbell GA, Sanoff S, Rosner MH. Care of the Undocumented Immigrant in the United States With ESRD. Am J Kidney Dis. 2010;55(1):181–191. doi:10.1053/j.ajkd.2009.06.039

23. United States Renal Data System. 2021 USRDS Annual Data Report: Epidemiology of kidney disease in the United States. Published online 2021.

24. Chapman CL, Hess HW, Lucas RAI, et al. Occupational heat exposure and the risk of chronic kidney disease of nontraditional origin in the United States. Am J Physiol-Regul Integr Comp Physiol. 2021;321(2):R141–R151. doi:10.1152/ajpregu.00103.2021

25. Harris Health System 2021-2025 Strategic Plan. https://www.harrishealth.org/SiteCollectionDocuments/strategic-plan.pdf

26. Profile of the Unauthorized Population – County Data (48201). migrationpolicy.org. Accessed April 11, 2023. https://www.migrationpolicy.org/data/unauthorized-immigrant-population/county/48201

27. Raghavan R. When access to chronic dialysis is limited: one center’s approach to emergent hemodialysis. Semin Dial. 2012;25(3):267–271. doi:10.1111/j.1525-139X.2012.01066.x

28. Raghavan R, Sheikh-Hamad D. Descriptive analysis of undocumented residents with ESRD in a public hospital system. Dial Transplant. 2011;40(2):78–81. doi:10.1002/dat.20535

29. Zhang HH, Cortés-Penfield NW, Mandayam S, et al. Dialysis Catheter–related Bloodstream Infections in Patients Receiving Hemodialysis on an Emergency-only Basis: A Retrospective Cohort Analysis. Clin Infect Dis Off Publ Infect Dis Soc Am. 2019;68(6):1011–1016. doi:10.1093/cid/ciy555

30. Wesseling C, Joode B van W de, Crowe J, et al. Mesoamerican nephropathy: geographical distribution and time trends of chronic kidney disease mortality between 1970 and 2012 in Costa Rica. Occup Environ Med. 2015;72(10):714–721. doi:10.1136/oemed-2014-102799

31. Gonzalez-Quiroz M, Smpokou ET, Silverwood RJ, et al. Decline in Kidney Function among Apparently Healthy Young Adults at Risk of Mesoamerican Nephropathy. J Am Soc Nephrol. 2018;29(8):2200–2212. doi:10.1681/ASN.2018020151

32. Raines N, González M, Wyatt C, et al. Risk factors for reduced glomerular filtration rate in a Nicaraguan community affected by Mesoamerican nephropathy. MEDICC Rev. 2014;16(2):16–22. doi:10.37757/MR2014.V16.N2.4

33. Marcantoni C, Ma LJ, Federspiel C, Fogo AB. Hypertensive nephrosclerosis in African Americans versus Caucasians. Kidney Int. 2002;62(1):172–180. doi:10.1046/j.1523-1755.2002.00420.x

34. Vikse BE, Aasarød K, Bostad L, Iversen BM. Clinical prognostic factors in biopsy-proven benign nephrosclerosis. Nephrol Dial Transplant. 2003;18(3):517–523. doi:10.1093/ndt/18.3.517

35. Wesseling C, Crowe J, Hogstedt C, Jakobsson K, Lucas R, Wegman DH. Resolving the Enigma of the Mesoamerican Nephropathy: A Research Workshop Summary. Am J Kidney Dis. 2014;63(3):396–404. doi:10.1053/j.ajkd.2013.08.014

36. R Core Team. R: A language and environment for statistical computing. Published online 2022. https://www.R-project.org/.

37. United States Renal Data System. 2016 USRDS Annual Data Report: Epidemiology of kidney disease in the United States. Published online 2016.

38. National Diabetes Statistics Report 2020. Estimates of diabetes and its burden in the United States. Published online 2020:32.

39. Schneiderman N, Llabre M, Cowie CC, et al. Prevalence of Diabetes Among Hispanics/Latinos From Diverse Backgrounds: The Hispanic Community Health Study/Study of Latinos (HCHS/SOL). Diabetes Care. 2014;37(8):2233–2239. doi:10.2337/dc13-2939

40. Fischer RSB, Vangala C, Mandayam S, et al. Clinical markers to predict progression from acute to chronic kidney disease in Mesoamerican nephropathy. Kidney Int. 2018;94(6):1205–1216. doi:10.1016/j.kint.2018.08.020

41. Trabanino RG, Aguilar R, Silva CR, Mercado MO, Merino RL. [End-stage renal disease among patients in a referral hospital in El Salvador]. Rev Panam Salud Publica Pan Am J Public Health. 2002;12(3):202–206. doi:10.1590/s1020-49892002000900009

42. Passel JS. Chapter 2: Birthplaces of U.S. Unauthorized Immigrants. Pew Research Center’s Hispanic Trends Project. November 18, 2014. Accessed November 4, 2022. https://www.pewresearch.org/hispanic/2014/11/18/chapter-2-birthplaces-of-u-s-unauthorized-immigrants/

43. Burden of Kidney Diseases – PAHO/WHO | Pan American Health Organization. Accessed July 25, 2022. https://www.paho.org/en/noncommunicable-diseases-and-mental-health/noncommunicable-diseases-and-mental-health-data-37

